# American older adults in COVID-19 Times:Vulnerability types, aging attitudes and emotional responses

**DOI:** 10.1101/2021.04.29.21256178

**Authors:** Mingqi Fu, Jing Guo, Xi Chen, Qilin Zhang

## Abstract

**Background:** The coronavirus disease aroused challenges to the emotional well-being of vulnerable older adults in hard-hit areas. This study investigates different vulnerability types among American older adults and how modes of vulnerability are associated with aging attitudes and emotional responses.

**Methods:** Using Latent Class Analysis, we investigated 2003 respondents aged over 50 from HRS. Hierarchical linear regressions with the affective profile as cluster identity were used to examine the relationship between vulnerability type and positive aging attitudes with positive and negative emotional responses.

**Results:** We detected three vulnerability types among American older adults: the slight vulnerability (72%), the healthcare use vulnerability (19%), and the dual vulnerabilities (9%). No significant difference in positive emotions was found between vulnerability types. However, more negative emotions were found among older adults with healthcare use vulnerability (B=0.746, SE=0.759) and dual vulnerabilities (B=1.186, SE=0.274) than those with slight vulnerability. Positive aging attitudes associate with more positive emotions (B=0.266, SE=0.017) but less negative emotions (B=-0.183, SE=0.016) and had significant moderation effects on the relationship between vulnerability types and negative emotional responses (B=-0.118, SE=0.045).

**Conclusion:** Older adults’ emotional well-being should not be neglected as they deserve the support of prevention and intervention strategies, in particular when they have vulnerabilities in healthcare use and financial sustainment. Female, non-white races, and those aged below 65, been uncoupled, less educated, and with ADL difficulties should prioritize.

## 1. Introduction

The coronavirus disease (COVID-19) became a global pandemic with substantial mental health impacts besides direct threats to individuals’ physical health (Pfefferbaum and North, 2020). Existing research reveals that negative emotionality was rising in many countries (Garrett, 2020). Individuals faced challenges to emotional well-being with the unease of pandemic threats, changes in routine, worries about financial loss, and loneliness from proposed distancing policies (Pfefferbaum and North, 2020). Whereas older adults generally have higher levels of overall emotional well-being (Carstensen *et al*., 2020), they might experience heightened negative emotions when faced with acute stresses such as the SARS outbreak (Lau *et al*., 2008). In the hard-hit United States, about one-half of the older adults reported stresses related to the disease, and a quarter of them developed passive mental health responses during the pandemic (Koma *et al*., 2020). According to the Strength and Vulnerability Integration Model (Charles, 2010), older adults have increased strengths in the successful use of attentional strategies, appraisals, and behaviors to regulate emotional experiences and vulnerability in modulating the high and sustained levels of physiological arousal. In the COVID-19 situation where stress is prolonged and inescapable, avoiding distress motion regulation skills is difficult. As emotions have a great potential in shaping life satisfaction and coping strategies (Fredrickson and Losada, 2005), a growing body of researches found impacts from physical inactivity, living alone, and perceived stresses were associate with emotional responses (Carstensen *et al*., 2020; Fingerman *et al*., 2021). Nevertheless, to our knowledge, a few studies examined emotional well-being with types of vulnerability.

Vulnerability refers to the inability of individuals to withstand the multiple effects of a hostile environment (Ahmad *et al*., 2020). The elderly, as a specific vulnerable group, has attracted substantial attention for their vulnerabilities relative to physical health indicators, cognition, and social orientation (Cesari *et al*., 2017; Morrow-Howell *et al*., 2020). Chambers proposes that vulnerability has two sides: an external side of risk, shock, and stress to which individuals are subjects. Also, an internal side means a lack of means to cope without damaging loss (Chambers, 2006). In the previous study, the vulnerability was assessed with the outcomes of a set of risks (Schröder-Butterfill and Marianti, 2006), with expressions primarily on trauma exposure, delayed healthcare use, financial hardships, and disrupted routine. As noted by the Sensitive and Resilience model (Charney, 2004), these dimensions face the most frequent disturbances during an upheaval, while older adults present prepositioning frailties on them (Morrow-Howell *et al*., 2020). However, the capability of older adults to withstand exposures and threats is highly variable, suggesting the vulnerability across older adults may differ qualitatively. One systematic review reveals that older adults with less physiological reserves were particularly vulnerable to physiologic changes, and those in disadvantaged social status were more sensitive to resource loss (Bayraktar and Dal Yilmaz, 2018). Personal protection, practical preparedness, and social preparedness combined to determine how an older adult becomes vulnerable to adverse events (Pérez-Galarce *et al*., 2017). However, we have limited evidence to date to understand the vulnerability across older adults in COVID-19 scenarios. Thus, this study examines the latent types of vulnerability among older adults in the COVID-19 context.

Previous studies proposed that vulnerabilities during adverse events might arouse negative responses (Lau *et al*., 2008; Knepple Carney *et al*., 2021), despite the emotional well-being is relatively stable and echoes affective profile (Garcia *et al*., 2014). However, prior researches reached no consensus regarding the association between vulnerability and positive emotional responses. In some studies, positive emotions decreased when the person had more or severer vulnerabilities (Thomas and Hasher, 2006; Ebner and Johnson, 2010). However, other studies found no significant reduction in positive emotions despite older adults were conceived as vulnerable (Carstensen and Mikels, 2005; Fuller and Huseth-Zosel, 2021). Differences in the interpretation of vulnerability, stress types, and cultures contributed to this inconsistency. As the Cognitive Appraisal Theory of Emotion reveals (Lazarus, 1991), positive emotional responses during an adverse event link with factual vulnerabilities and ruminations afterward. In unintentional situations, positive internal schema and positive emotions among older adults would sustain when they attributed vulnerabilities to external factors or conceived them as a shared risk for the whole society. Alternatively, considering the vulnerabilities as personal inability may lead to self-depreciation and expel positive emotions. During the COVID-19, how older adults conceive vulnerabilities may be differential, as risks from the pandemic are commonly shared by nearly all communities and groups (Calderón-Larrañaga *et al*., 2020). Thus, in the current study, we examine the relationships of vulnerabilities with both positive and negative emotional responses, thereby evaluating the emotional well-being of older adults amidst COVID-19 threats.

Moreover, aging attitude as a cognitive pattern might moderate the relationship between vulnerability and emotional responses. In line with the Cognitive Vulnerability-Stress Model (Beck, 2002), a sizable of studies concurred that older adults with positive aging attitudes would have better subjective well-being during adverse events (Reichstadt *et al*., 2007; Suh *et al*., 2012). When confronted with factual vulnerabilities, positive aging attitudes improve emotional well-being via emotional and informational processing. On the one hand, self-esteem from a sense of self-identity serves as a shield against initial negative reactions, which is helpful to restrain stress-diathesis and have successful use of emotion regulation strategies (Hooley and Gotlib, 2000). On the other hand, positive attitudes towards aging offer older persons better preparedness to capitalize on the strengths of aging by effectively selecting information that increases positive effects (Ayalon, 2020). Also, individuals holding positive views on aging are less likely to consider older adults’ vulnerabilities as social discrimination, thus conducting negative ruminations to a lesser extent. However, a few studies explored the differences between vulnerability types in the relationship of vulnerability, aging attitudes, and emotional well-being to our knowledge. Theoretically, older adults with fewer or mild vulnerabilities may have greater self-esteem together with confidence to overcome the pandemic after comparing with their severely damaged counterparts. Such a sense of capability is intrinsically inherent with positive aging attitudes and might amplify their protective effects on emotional responses (Chen *et al*., 2018). By contrast, another study reveals that positive aging attitudes are of utmost importance for older adults whose substantial vulnerabilities had threatened the pre-existing positive cognitions (Bellingtier and Neupert, 2018). Establishing the interaction of vulnerability and aging attitudes is a necessary step in proposing intervention strategies regarding emotional well-being.

This study investigates the latent vulnerability types in American older adults in the face of COVID-19 threats and examines the relationships between vulnerability type, aging attitudes, and emotional responses. Our first hypothesis concerns the latent vulnerability types with investigations on pandemic exposure, healthcare use, financial resilience, and routine sustainment. This hypothesis was exploratory, as the vulnerability of older adults may be represented by several distinct types, with specific sociodemographic characteristics for each type. Second, we propose that greater vulnerabilities are associated with more negative emotional responses. In contrast, the relationship between vulnerability and positive emotional responses might be cross-current or nonsignificant. Last, we hypotheses that vulnerable older adults would have better emotional well-being when they had positive rather than negative aging attitudes. Moreover, salutary effects of positive cognition on emotional responses might be differential across vulnerability types.

## 2. Methods

### Study design and data collection

Health and Retirement Survey (HRS) is a national longitudinal study of older Americans’ health and economic situation. Data in this study were from the 2020 HRS COVID-19 Project (Early, version 1.0), with predetermined affective profiles from HRS 2002-2018 waves. The COVID-19 module is being administrated 50% random subsample of households initially assigned to enhance face-to-face interviewing. This 50% random subsample was further split into two random samples, while the first one was released to fieldwork on June 11, 2020, and the second one on September 24, 2020. Information in this study is from the first random sample of 3266 respondents, accounting for approximately 25% of the original HRS sample. Due to lockdowns in the pandemic, the COVID-19 Project was conducted via telephone, with a response rate of 62%. To adjust for selections and non-response into the data, the researchers have created preliminary weights for respondents. Detailed information is available at the official website https://hrsonline.isr.umich.edu/. After excluding persons aged below 50 years old or who did not report psychological evaluations, this study included a total of 2003 older adults, accounting for 61.3% of the surveyed sample.

Collection and production of HRS data comply with the requirements of the University of Michigan’s Institutional Review Board (IRB). All respondents gave verbal consent to this survey.

## 3. Measures

### Outcome Variable

**Emotional responses** in this study were measured with the International Positive and Negative Affect Schedule Short-form (I-PANAS-SF, Thompson, 2007), estimating which degree of positive and negative affectivities for individuals experienced in the past month. Ten items in this scale were retrieved from the original 20 PANAS (Watson *et al*., 1988) items pool. Five positive emotional responses are active, determined, attentive, inspired, and alert, whereas five negative responses including afraid, nervous, upset, hostile, and ashamed. Older adults were invited to rate these emotional responses on a 5-point scale according to the extent to which each describes the way they have felt, while higher scores are referring to more intensive affectivities. The I-PANAS-SF is psychometrically acceptable across cultures (Karim *et al*., 2011). In the current study, the Cronbach’s α for positive and negative emotionality was 0.811, 0.776, respectively.

### Independent Variables

**Aging Attitudes** were examined with a brief five-item unidimensional measure that compromises the Attitudes toward own aging (ATOA) dimensions of the Philadelphia Geriatric Center (PGC) Morale Scale (Lawton, 1975). Items from the ATOA measure including “I have much pep as I did last year”, “I am as happy now as when I was younger”, “Things keep getting worse as I get older”, “The older I get, the more useless I feel”, and “As I get older, things are better than I thought they would be”. A 6-point response scale was used in evaluate the degree of each item. When negative items were reverse-scored, this scale captures older adults’ global positive evaluation of their aging process (Kim *et al*., 2014). The Cronbach’s α for the scale was 0.773 in this study.

**The Vulnerability** of older adults focuses on 16 kinds of difficulties they might experience in the COVID-19 era (Items displayed in Table 1). Individuals reported if they had experienced each of the 16 items, with 0 denotes for no, and one represents yes.

**Table 1.**
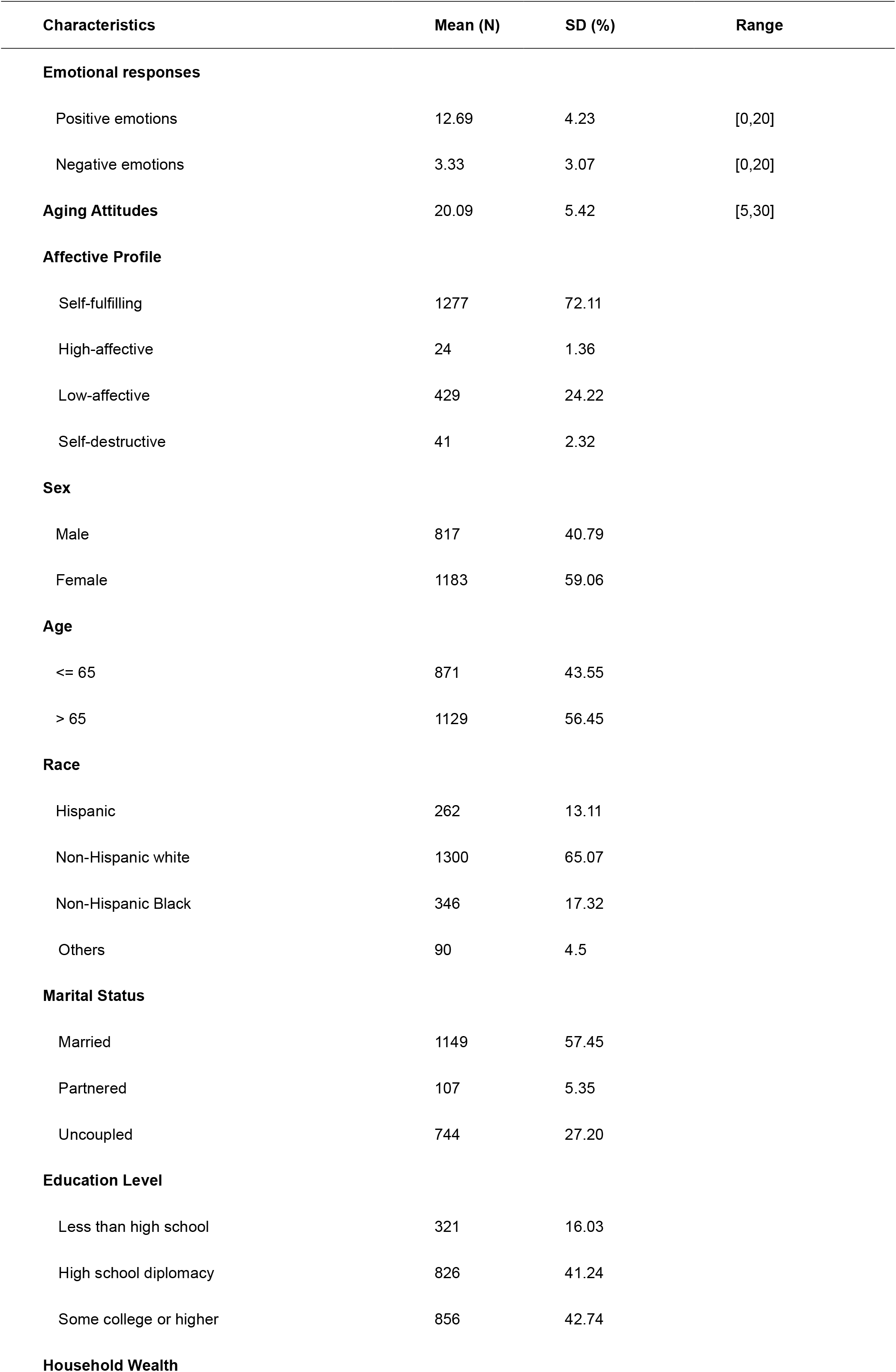
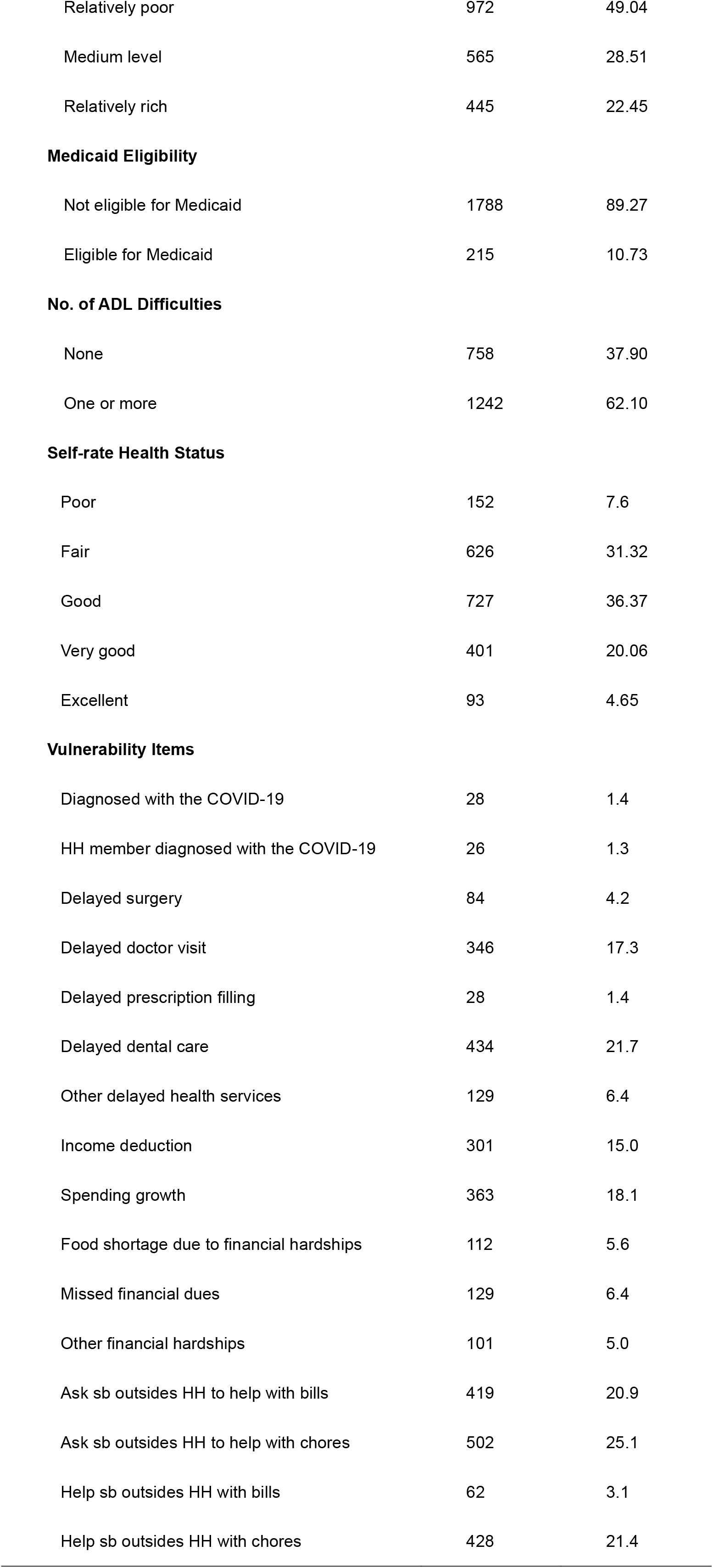
Descriptive analysis of the sample (N=2003).

| Characteristics | Mean (N) | SD (%) | Range |
| --- | --- | --- | --- |
| **Emotional responses** |  |  |  |
| Positive emotions | 12.69 | 4.23 | [0,20] |
| Negative emotions | 3.33 | 3.07 | [0,20] |
| **Aging Attitudes** | 20.09 | 5.42 | [5,30] |
| **Affective Profile** |  |  |  |
| Self-fulfilling | 1277 | 72.11 |  |
| High-affective | 24 | 1.36 |  |
| Low-affective | 429 | 24.22 |  |
| Self-destructive | 41 | 2.32 |  |
| **Sex** |  |  |  |
| Male | 817 | 40.79 |  |
| Female | 1183 | 59.06 |  |
| **Age** |  |  |  |
| <= 65 | 871 | 43.55 |  |
| > 65 | 1129 | 56.45 |  |
| **Race** |  |  |  |
| Hispanic | 262 | 13.11 |  |
| Non-Hispanic white | 1300 | 65.07 |  |
| Non-Hispanic Black | 346 | 17.32 |  |
| Others | 90 | 4.5 |  |
| **Marital Status** |  |  |  |
| Married | 1149 | 57.45 |  |
| Partnered | 107 | 5.35 |  |
| Uncoupled | 744 | 27.20 |  |
| **Education Level** |  |  |  |
| Less than high school | 321 | 16.03 |  |
| High school diplomacy | 826 | 41.24 |  |
| Some college or higher | 856 | 42.74 |  |
| **Household Wealth** |  |  |  |

|  |  |  |
| --- | --- | --- |
| Relatively poor | 972 | 49.04 |
| Medium level | 565 | 28.51 |
| Relatively rich | 445 | 22.45 |

**Medicaid Eligibility**
|  |  |  |
| --- | --- | --- |
| Not eligible for Medicaid | 1788 | 89.27 |
| Eligible for Medicaid | 215 | 10.73 |

**No. of ADL Difficulties**
|  |  |  |
| --- | --- | --- |
| None | 758 | 37.90 |
| One or more | 1242 | 62.10 |

**Self-rate Health Status**
|  |  |  |
| --- | --- | --- |
| Poor | 152 | 7.6 |
| Fair | 626 | 31.32 |
| Good | 727 | 36.37 |
| Very good | 401 | 20.06 |
| Excellent | 93 | 4.65 |

**Vulnerability Items**
|  |  |  |
| --- | --- | --- |
| Diagnosed with the COVID-19 | 28 | 1.4 |
| HH member diagnosed with the COVID-19 | 26 | 1.3 |
| Delayed surgery | 84 | 4.2 |
| Delayed doctor visit | 346 | 17.3 |
| Delayed prescription filling | 28 | 1.4 |
| Delayed dental care | 434 | 21.7 |
| Other delayed health services | 129 | 6.4 |
| Income deduction | 301 | 15.0 |
| Spending growth | 363 | 18.1 |
| Food shortage due to financial hardships | 112 | 5.6 |
| Missed financial dues | 129 | 6.4 |
| Other financial hardships | 101 | 5.0 |
| Ask sb outsides HH to help with bills | 419 | 20.9 |
| Ask sb outsides HH to help with chores | 502 | 25.1 |
| Help sb outsides HH with bills | 62 | 3.1 |
| Help sb outsides HH with chores | 428 | 21.4 |

### Covariates

**Affective profile** was calculated from the mean score of emotional responses reported throughout 2002-2018 HRS waves and was believed to have an anchoring effect on emotional responses during the COVID-19 pandemic. In line with previous studies, we adopted a cutoff point at 53.2% for positive emotions and 48.9% for negative emotions to explore the orthogonal structure of emotions (Garcia *et al*., 2014). Four terms were used to distinguish affective profiles in this study: self-fulfilling profile (high scores in PA but low scores in NA); low affective profile (low scores both in PA and NA); high affective profile (high scores both in PA and NA), and self-destructive profile (low scores in PA but high scores in NA).

Other covariates in the current study including sex (male/female), age (≤65/ > 65), race (Hispanic/Non-Hispanic white/Non-Hispanic Black/Others), marital status (married/partnered/uncoupled), educational level (less than high school/high school diplomacy/some college or above), household wealth (relatively poor/mid-level/rich), Medicaid eligibility (yes/no), number of difficulties in activities of daily life (none/one and more) and self-reported health status (poor/fair/good/very good/excellent). These variables are associated with subjective well-being among older adults in the United States (de Main and Xie, 2020).

### Statistical analyses

Descriptive analyses were conducted for the outcome variable, independent variables, and all covariates. We used Latent class analysis (LCA) to identify unobserved clusters of individuals that respond to measured vulnerability expressions with a similar pattern. In this stage, robust maximum likelihood (MLR) estimators were adopted. Indicators such as Akaike Information Criterion (AIC), Bayesian Information Criterion (BIC), sample size-adjusted BIC (ssaBIC), entropy, values of the Lo-Mendell-Rueben Test (LMRT), and the Bootstrap Likelihood Ratio Test (BLRT) were used for model selection. In the next step, class membership was assigned to each individual based on the probability, while uncertainty with this assignment was controlled after adjusting the classification uncertainty rates (Asparouhov and Muthén, 2014). Meanwhile, descriptive analyses on socioeconomic and health-related variables for each vulnerability type, along with the differences between classes (evaluated via Chi-square or F tests), were conducted. Lastly, Hierarchy linear regressions with emotional responses as outcome variables and affective profile as the Level-2 identity were conducted. Associations between aging attitudes, vulnerability types, and emotional experiences were explored through nested models. Latent class analysis was conducted via Mplus Version 7, and regressions were conducted with Stata Version 14.

## 4. Results

**Table 1** reports descriptive characteristics of listed variables in this study. Among 2003 respondents, female (N=1183, 59.06%) and those over 65 years old (N=1129, 56.45%) accounted for the majority. Additionally, more than half of the respondents were Non-Hispanic white (N=1300, 65.07%), been married or coupled (N=1256, 62.80%), and had high school education or above (N=1682, 83.98%). Although 62.1% (N=1242) of the elderly had ADL difficulty(s), the proportion for a self-reported poor or fair health status was only 38.92% (N=778). In line with the high proportion of self-fulfilling affective profile (N=1277, 72.11%), older adults in this study reported averagely higher positive (Mean=12.69, SD=4.23, range 0-20) versus negative scores (Mean=3.33, SD=3.07, range 0-20) in emotional responses. Regarding vulnerability items, unable to do chores (N=502, 25.10%) or pay the bills (N=419, 20.9%) by themselves, had to help others with the chores (N=428, 21.4%), and had delayed dental care (N=434, 21.7%) were the most frequent ones. The mean score of positive aging attitudes was 20.09 (SD=5.42, range 5-30), and other details are shown in **Table** 1.

Latent Class Analysis reveals that older adults in the current study developed three distinct patterns of vulnerability when confronted with the COVID-19 threat. Accordingly, older adults distributed unfairly across vulnerability types, as the proportion in three classes was 19%, 9%, and 72%, respectively. (see **Table S1**)

**Figure 1** presents the detected vulnerability types based on the estimated probability of respondents from each latent class answered yes to vulnerability items. The solid black line refers to older adults who demonstrated slight vulnerabilities during the COVID-19 pandemic. Apart from several experiences commonly witnessed by populations, such as income deduction and spending growth, older adults in this group are prone to experience a certain degree of inconveniences regarding chores and bills. About 72% of older adults in this study had slight vulnerabilities in sustaining daily routine and could be comparatively healthier and more capable than two other types. Besides, about one in five of the older adults had a vulnerability in healthcare use. Comparatively, they had intense demands in surgery, doctor visits, dental care et al., but failed to accommodate due to lockdowns in COVID-19. Noteworthy, another 9% of older adults were comprehensively vulnerable when confronted with the COVID-19 threat. Besides elevated risks in life inconveniences and pandemic exposure, persons in this group are prone to experience healthcare shortages and economic hardships. A 20% to 50% probability of experiencing delayed survey, doctor visits, and dental care, along with income deduction, spending growth, food shortage, and financial due miss, went to older adults with dual vulnerabilities. Details could be seen in **Figure 1**.

**Figure 1.**
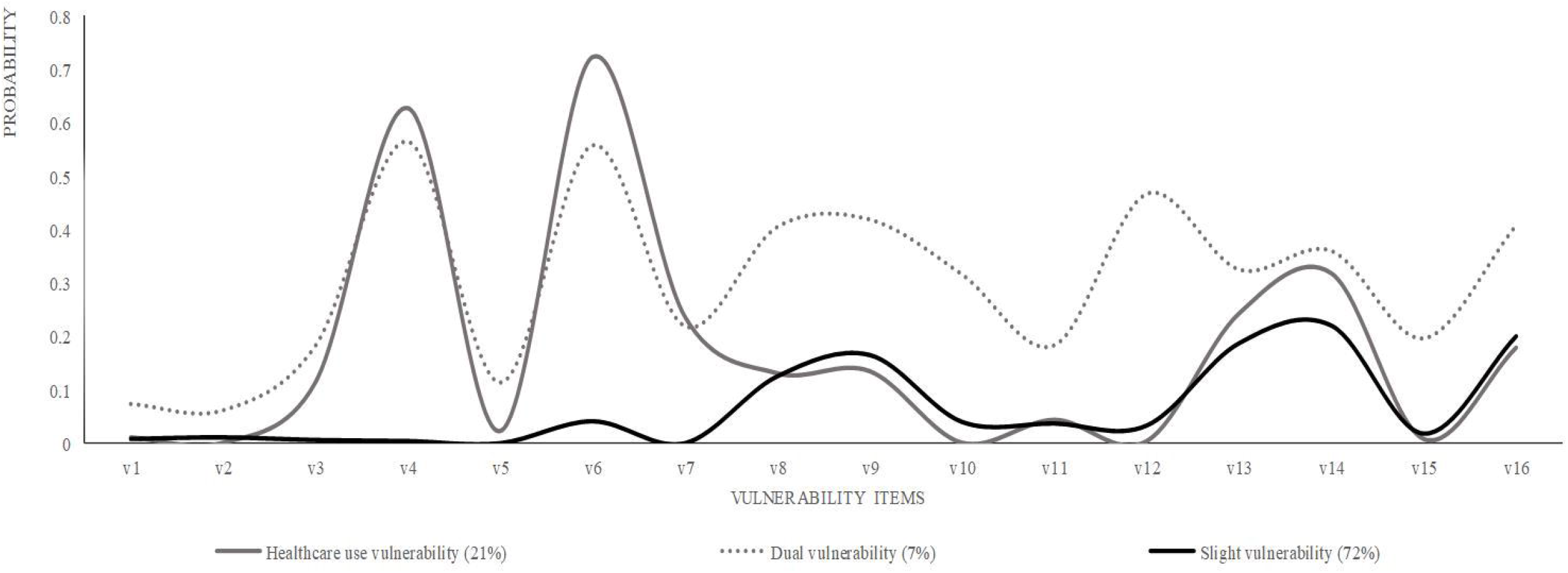
Vulnerability Groups Detected among Older Adults in the United States during the COVID-19 pandemic. **Notes:** vl to vl6 are items used to measure the vulnerability of older adults during the pandemic. **v1** Been diagnosed with the COVID-19; **v2** Has household member been diagnosed with the COVID-19; **v3** Had delayed surgery; **v4** Had delayed doctor visit; **v5** Had delayed prescription filling; **v6** Had delayed dental care; **v7** Had other delayed healthcare services; **v8** Experienced income deduction; **v9** Experienced spending growth; **v10** Had food shortage due to financial hardships; **v11** Missed financial dues; **v12** Had other financial hardships; **v13** Ever asked help from somebody outsides household to pay bills during the pandemic; **v14** Ever asked help from somebody outsides household to do chores during the pandemic; **v15** Helped somebody outsides household to pay their bills during the pandemic; **v16** Helped somebody outsides household to do chores during the pandemic.

**Table 2** further demonstrates each vulnerability type’s sociodemographic and health-related characteristics and identifies the differences between them. As noted in Table 3, there was no significant difference in affective profiles between individuals from different vulnerability types. Comparatively, the female was prone to have either healthcare use vulnerability or dual vulnerabilities other than slight vulnerability, whereas older adults aged over 65 were more likely to experience slight vulnerability. The proportion of Non-Hispanic white presented significant differences between vulnerability types, which was highest in healthcare use vulnerability (72.47%) and lowest in the dual vulnerability type (39.13%). Besides, older adults with less education, had ADL difficulties, and been uncoupled were most likely to have dual vulnerabilities. Details are presented in **Table 2**.

**Table 2.** Distribution of Level-2 Affective Profiles and Level-1 Characteristics across the Identified Latent Vulnerability Groups (N=2003).

| Characteristics | Class Description |  |  | Overall chi-square/F Test |  |
| --- | --- | --- | --- | --- | --- |
|  | Slight Vulnerability | Healthcare Use Vulnerability | Dual Vulnerability | Chi-square/F | P value |
| N (%) | 1439(71.84) | 426(21.27) | 138(6.89) |  |  |
| **Level-2 Affective Profile** |  |  |  | 1.25 | 0.29 |
| Self-fulfilling % | 73.49 | 72.70 | 54.78 |  |  |
| High affective % | 1.25 | 1.05 | 3.48 |  |  |
| Low affective % | 23.69 | 23.88 | 31.30 |  |  |
| Self-destructive % | 1.57 | 2.36 | 10.43 |  |  |
| **Level-1 Features** |  |  |  |  |  |
| Female % | 57.31 | 63.62 | 64.49 | 6.33** | 0.01 a, b |
| Age > 65 % | 60.17 | 49.77 | 38.41 | 20.34*** | <0.001 a, b |
| Non-Hispanic white % | 65.37 | 72.47 | 39.13 | 14.96*** | <0.001 a, b, c |
| Uncoupled % | 36.14 | 35.92 | 52.17 | 4.05* | 0.02 b, c |
| Less than high school % | 17.10 | 10.56 | 21.74 | 10.81*** | <0.001 a, c |
| Relatively poor % | 49.68 | 47.17 | 48.15 | 1.48 | 0.23 |
| Eligible for Medicaid % | 10.70 | 7.04 | 22.46 | 3.96 | 0.05 |
| With ADL Difficulties % | 60.79 | 60.80 | 79.71 | 9.85*** | <0.001 b, c |
| Poor health status % | 7.87 | 7.06 | 6.52 | 0.85 | 0.49 |

**Notes.**
Significant difference by class (P<0.05 with multinominal regression detecting partial relationship between group membership and each variable after controlling for covariates): a: Slight VUL vs Healthcare use VUL; b: Slight VUL vs Dual VUL; c: Healthcare use VUL vs Dual VUL.
P<0.05 \*, P<0.01 \*\*, P<0.001 \*\*\*.

**Table 3.** Mixed Effects ML Regressions of the Relationship between Aging Attitudes, Vulnerability Group and Positive Emotional Responses among Older Adults (Level-2 Identity: Affective Profile, N=2003).

| Variables |  | Model 1 |  | Model 2 |  | Model 3 |  |
| --- | --- | --- | --- | --- | --- | --- | --- |
| Fixed-effects |  | B | S.E. | B | S.E. | B | S.E. |
| Constant |  | 12.614 | 1.077 | 6.538 | 0.988 | 6.393 | 0.998 |
| **Gender** (ref: Male) | Female | 0.414* | 0.176 | 0.254 | 0.167 | 0.250 | 0.167 |
| **Marital Status** (ref: Married) | Partnered | -0.002 | 0.397 | 0.051 | 0.375 | 0.076 | 0.375 |
|  | Uncoupled | -0.462* | 0.192 | -0.338 | 0.182 | -0.356 | 0.182 |
| **Age** (ref: <=65) | >65 | -0.121 | 0.176 | 0.078 | 0.167 | 0.119 | 0.168 |
| **Education** (ref: < High School) | High school diplomacy | 0.442 | 0.258 | 0.438 | 0.244 | 0.425 | 0.244 |
|  | Some college or higher | 0.683* | 0.268 | 0.687** | 0.254 | 0.661** | 0.254 |
| **Race** (ref: Non-Hispanic White) | Hispanic | 0.104 | 0.278 | -0.204 | 0.264 | -0.246 | 0.265 |
|  | Non-Hispanic Black | 0.754*** | 0.234 | 0.246 | 0.225 | 0.224 | 0.226 |
|  | Others | 0.706 | 0.411 | 0.292 | 0.391 | 0.282 | 0.391 |
| **ADL Difficulties** (ref: None) | One or more | -0.746*** | 0.191 | -0.420* | 0.183 | -0.436* | 0.183 |
| **Self-rate Health** (ref: Poor) | Fair | -1.006** | 0.332 | -0.551 | 0.316 | -0.546 | 0.316 |
|  | Good | -2.085*** | 0.342 | -1.178*** | 0.329 | -1.176*** | 0.329 |
|  | Very good | -2.691*** | 0.384 | -1.098** | 0.377 | -1.097** | 0.377 |
|  | Excellent | -3.682*** | 0.510 | -1.284** | 0.506 | -1.365** | 0.508 |
| **Medicaid Eligibility** (ref: No) | Eligible | -0.172 | 0.297 | -0.080 | 0.284 | -0.082 | 0.284 |
| **Household Wealth** (ref: Poor) | Medium level | 0.608** | 0.202 | 0.611** | 0.192 | 0.605** | 0.192 |
|  | Relatively rich | 0.622** | 0.222 | 0.616** | 0.211 | 0.617** | 0.211 |
| **Aging Attitudes** (AA) |  |  |  | 0.263*** | 0.017 | 0.266*** | 0.017 |
| **VUL Group** (ref: Slight VUL) | Healthcare use VUL |  |  |  |  | 0.190 | 0.191 |
|  | Dual VUL |  |  |  |  | 0.591 | 0.328 |
| **Variance components** |  |  |  |  |  |  |  |
| var (_cons) |  | 1.936 | 0.712 | 1.579 | 0.586 | 1.607 | 0.597 |
| var (Residual) |  | 3.379 | 0.057 | 3.167 | 0.054 | 3.165 | 0.054 |
| **Obs** |  | 1752 |  | 1709 |  | 1709 |  |
| **Chi-square** |  | 244.35*** |  | 504.69*** |  | 509.58*** |  |
| **Ho: OLS versus HLM** |  | <0.001 |  | <0.001 |  | <0.001 |  |

**Notes:** AA: Aging Attitudes, VUL: Vulnerability; \* P<0.05; \*\* P<0.01; \*\*\* P<0.001.

In **Table 3**, the relationships between vulnerability type, aging attitudes, and positive emotional responses during the COVID-19 pandemic are presented. As affective profile was hypothesized with an anchoring effect on emotional responses, this study used the affective profile as a Level-2 identity. Socioeconomic and health-related variables entered on the first step, and aging attitudes were entered on the second step to predict positive emotional responses. In the second model, positive aging attitudes presented a significant association with positive emotional responses towards the pandemic (B=0.263, SE=0.017), also explained an additional 6.27% of the variability in positive emotions over and above socioeconomic and health-related factors. However, the third model indicated that no significant differences in positive emotional responses existed between vulnerability types. With aging attitudes and vulnerability types controlled, had some college or higher education (B=0.661, SE=0.254) and lived with better economic situations (B=0.605, SE=0.192 for mid-level; B=0.617, SE=0.211 for rich) was associated with more positive emotional responses. On the contrary, persons who had ADL difficulties (B=-0.436, SE=0.183) and perceived themselves as good or better health status were less likely to have positive emotions than their counterparts. See more details in **Table 3**.

**Table 4** focuses on negative emotional responses and explores their relationship with aging attitudes and vulnerability types. A set of hierarchical linear regressions was conducted with socioeconomic and health-related variables entered in the first step, and aging attitudes entered the second step to predict negative emotional responses. Positive aging attitudes explained 5.16% of the variance in negative emotions and presented a significant relationship with these responses (B=-0.194, SE=0.014). The vulnerability type entered in the third model, revealing that older adults who had healthcare use vulnerability (B=0.476, SE=0.159) and dual vulnerabilities (B=1.186, SE=0.274) were prone to develop more passive emotions than their slightly vulnerable counterparts. Moreover, there was a significant moderation effect of aging attitudes and vulnerability type on negative emotional responses. Positive aging attitudes are critical for persons being dual vulnerable, as negative emotions reduced faster in older adults with dual vulnerabilities than the slight vulnerable counterparts when aging attitudes improved (B=-0.118, SE=0.045). With aging attitudes and vulnerability type controlled, female (B=0.542, SE=0.140) was more likely to have negative emotions than male counterparts, whereas having Medicaid (B=-0.530, SE=0.253) and being uncoupled (B=-0.336, SE=0.152) associated with fewer negative emotional responses.

**Table 4.** Mixed Effects ML Regressions of the Relationship between Aging Attitudes, Vulnerability Type and Negative Emotional Responses among Older Adults (Level-2 Identity: Affective Profile, N=2003).

| Variables |  | Model 1 |  | Model 2 |  | Model 3 |  | Model 4 |  |
| --- | --- | --- | --- | --- | --- | --- | --- | --- | --- |
| Fixed-effects |  | B | S.E. | B | S.E. | B | S.E. | B | S.E. |
| Constant |  | 2.930 | 0.871 | 7.762 | 0.951 | 8.513 | 0.988 | 8.204 | 0.989 |
| **Gender** (ref: Male) | Female | 0.493*** | 0.146 | 0.564*** | 0.140 | 0.553*** | 0.140 | 0.542*** | 0.140 |
| **Marital Status**<br>(ref: Married) | Partnered | -0.231 | 0.329 | -0.281 | 0.314 | -0.274 | 0.314 | -0.268 | 0.313 |
|  | Uncoupled | -0.283 | 0.158 | -0.380* | 0.152 | -0.371* | 0.152 | -0.366* | 0.152 |
| **Age** (ref: <=65) | >65 | -0.616*** | 0.147 | -0.749*** | 0.141 | -0.735*** | 0.141 | -0.727 | 0.141 |
| **Education**<br>(ref: < High School) | High school diplomacy | -0.198 | 0.214 | -0.162 | 0.205 | -0.154 | 0.205 | -0.114 | 0.205 |
|  | Some college or higher | 0.166 | 0.222 | 0.191 | 0.213 | 0.191 | 0.213 | 0.225 | 0.213 |
| **Race**<br>(ref: Non-Hispanic White) | Hispanic | -0.006 | 0.231 | 0.323 | 0.223 | 0.332 | 0.223 | 0.374 | 0.224 |
|  | Non-Hispanic Black | -0.110 | 0.193 | 0.309 | 0.188 | 0.324 | 0.189 | 0.336 | 0.189 |
|  | Others | -0.805* | 0.340 | -0.458 | 0.328 | -0.433 | 0.327 | -0.439 | 0.326 |
| **ADL Difficulties** (ref: No) | One or more | 0.302 | 0.158 | -0.002 | 0.153 | 0.001 | 0.153 | 0.009 | 0.153 |
| **Self-rate Health** (ref: Poor) | Fair | 0.560* | 0.272 | 0.196 | 0.262 | 0.196 | 0.262 | 0.197 | 0.262 |
|  | Good | 0.740** | 0.280 | 0.109 | 0.273 | 0.105 | 0.273 | 0.128 | 0.273 |
|  | Very good | 1.488*** | 0.316 | 0.380 | 0.315 | 0.375 | 0.314 | 0.402 | 0.314 |
|  | Excellent | 2.347*** | 0.420 | 0.763 | 0.422 | 0.789 | 0.422 | 0.706 | 0.422 |
| **Medicaid** (ref: No) | Eligible | -0.433 | 0.244 | -0.530* | 0.236 | -0.525* | 0.236 | -0.530* | 0.235 |
| **Household Wealth**<br>(ref: Poor) | Medium level | -0.017 | 0.167 | -0.038 | 0.161 | -0.045 | 0.161 | -0.055 | 0.161 |
|  | Relatively rich | 0.058 | 0.183 | 0.003 | 0.176 | -0.005 | 0.176 | -0.003 | 0.175 |
| **Aging Attitudes** (AA) |  |  |  | -0.194*** | 0.014 | -0.194*** | 0.014 | -0.183*** | 0.016 |
| **VUL Group**<br>(ref: Slight VUL) | Healthcare use VUL |  |  |  |  | 0.746*** | 0.159 | 0.948 | 0.620 |
|  | Dual VUL |  |  |  |  | 1.186*** | 0.274 | 3.245*** | 0.841 |
| **Inter** (ref: Slight VUI *AA) | Healthcare VUL * AA |  |  |  |  |  |  | -0.010 | 0.030 |
|  | Dual VUL * AA |  |  |  |  |  |  | -0.118** | 0.045 |
| **Variance components** |  |  |  |  |  |  |  |  |  |
| var (_cons) |  | 1.454 | 0.715 | 1.528 | 0.726 | 1.702 | 0.737 | 1.673 | 0.628 |
| var (Residual) |  | 2.754 | 0.047 | 2.612 | 0.045 | 2.609 | 0.045 | 2.604 | 0.045 |
| **Obs** |  | 1709 |  | 1668 |  | 1668 |  | 1668 |  |
| **Chi-square** |  | 105.29*** |  | 299.05*** |  | 374.92*** |  | 383.17*** |  |
| **Ho: OLS versus HLM** |  | <0.001 |  | <0.001 |  | <0.001 |  | <0.001 |  |

**Notes:** AA: Aging Attitudes, VUL: Vulnerability; \* P<0.05; \*\* P<0.01; \*\*\* P<0.001.

## 5. Discussion

The COVID-19 pandemic has put older adults at increased risks of life disruptions and negative emotional responses. This study reveals that older adults aging in the COVID times had three latent classes regarding their vulnerability. The proportion of older persons as slightly vulnerable, the healthcare uses vulnerable, and the dual vulnerable was 72%, 19%, and 9%, respectively. With the affective profile as a cluster factor, vulnerability presented no significant association with positive emotions. However, older adults who had healthcare uses or dual vulnerabilities developed more negative emotions than their slightly vulnerable counterparts. Positive aging attitudes associated with better emotional well-being generally and moderated the relationship between vulnerability types and negative emotional responses. Findings in this study provide a better understanding of the vulnerability of aging in the COVID-19 and shed light on emotional well-being promotion strategies in pandemic contexts.

First, we propose that older adults in the United States have significant but differential types of vulnerabilities during the COVID-19 pandemic and 28% of them experienced delayed healthcare needs solely or together with financial hardships. Due to the non-availability of transport and cancellation of appointments, the delayed needs of healthcare were prevalent during the pandemic, especially in hard-hit areas where health resources were massively reassigned for pandemic control (Cudjoe and Kotwal, 2020). Compared with younger adults, older adults are more vulnerable given their additional physical illness and unfamiliarity with telehealth applications. For example, one contemporaneous study conducted among U.S. adults proposed a 46.6% prevalence of delayed dental care during the pandemic (Kranz *et al*., 2021). In contrast, the probability for such delay was about 55% to 70% among older adults in the current study. Another report claimed that doctor visits for U.S. adults from June to September in 2020 were about to 86% of the pre-pandemic level (IQVIA, 2020). However, more than half of the older adults reported delayed doctor visits in this study. Noteworthy, nearly one in ten older persons had dual vulnerabilities in financial hardships and delayed healthcare use, which is consistent with the previous estimation on the poverty rate of American elderly (Semega *et al*., 2019). As half of the older adults live without emergency savings (Catherine S. Harvey, 2019), and 25% of them depend largely or exclusively on social security benefits (Dushi *et al*., 2017), it is worrisome that more individuals would fell below the threshold of poverty if pension funding had shortfalls. Thus, we urge more attention to older adults’ vulnerabilities regarding healthcare use and financial sustainment. Public health programs in pandemics should consider the necessity of timely healthcare delivery and financial support to the elderly.

Additionally, females, BLOAC (black, Latinx, and older adults of color), and those aged below 65 years old, been uncoupled, less educated, and with ADL difficulties were more likely to be severely vulnerable. In line with previous researches (Harman, 2016; Wenham *et al*., 2020), this study proposes that women experienced greater risks in delayed healthcare use and financial hardships during pandemics. On the one hand, women take more care of their health than their male counterparts in general but are more likely to skip medical appointments due to greater fear of infection and social inequalities such as inadequate health coverage (Allen *et al*., 2020). On the other hand, women are less likely than men to have power in decision-making when emergencies occurred, and their needs in healthcare and economic concerns were largely unmet (Harman, 2016). Besides, consistent with previous studies (Black *et al*., 2019; Kranz *et al*., 2021), BLOAC and those uncoupled, less educated, and with ADL difficulties have elevated vulnerabilities given their greater health and financial risks versus limited capabilities. However, it is counterintuitive that the young-old developed more vulnerabilities than the old-old counterparts, as prior studies prone to take age as a risk factor of vulnerability (Cesari *et al*., 2017; Morrow-Howell *et al*., 2020). Two possible explanations might contribute to this inconsistency. On the financial vulnerability, while older adults aged over 65 have a stable income from social security benefits, most of the young-old are still active in the labor market. During the economic recession in pandemics, the young-old had difficulty sustaining their income and reentering the workforce (Pfefferbaum and North, 2020), thereby more chance to experience financial hardships. On the other hand, the old-old may have developed greater tolerance to their illness for healthcare use vulnerability and more familiar with telemedicine visits than the young-old do. As one prior study supports, older adults aged 55-65 had fewer telemedicine visits than their older counterparts (Eberly *et al*., 2020). Findings on each vulnerability type’s sociodemographic characteristics help target the older adults severely damaged during the pandemic.

Third, this study reveals that the vulnerability of older adults presents no significant relationship with positive emotions but associates with more negative emotional responses during the pandemic. Previous studies claimed that more significant vulnerabilities would lead to a ruined sense of self-continuity and additional stress from traumatic loss, reducing positive emotions in adults (Pallant and Lae, 2002; Fogel, 2009). However, this study found no significantly reduced positive emotions among older adults with vulnerabilities. On the one hand, the sustainment of positive emotions might result from conscious efforts of which older adults perceived their vulnerabilities as prevalent and occasional, thus reducing negative impacts of vulnerability on subjective well-being. On the other hand, there might also exist an autonomous protective mechanism regarding positive emotions in older adults. Given that over 70% of the respondents reported the self-fulfilling affective profile, it is reasonable to assume that older adults in this study developed socioemotional selectivity and preferential attention to positive stimuli over negative information (Reed *et al*., 2014). The intrinsic motivation for emotionally meaningful goals has stronger associations with positive emotional regulations than negative emotional responses (Vandercammen *et al*., 2014). Nevertheless, this study suggests that negative reactions are more sensitive to external changes during adverse events. In accordance, one previous study found that when older adults were engaged in interactions with a hostile environment, they would experience the same levels of negative reactivities as the younger people (Charles *et al*., 2009). Another study analyzes with a situation selection perspective (Birditt, 2014). It proposes that older adults reported lesser negative responses to mildly distressing events that they could largely adapt to, such as disrupted life routines. However, when required to engage in unavoidable but stressful events, more negative emotions would onset among older adults (Birditt, 2014). Thus, with the aim of emotional well-being promotion, this study urges that prevention and intervention on negative reactivities are of utmost importance.

Last, we propose that positive aging attitudes benefit older adults’ emotional well-being in the COVID-19 context, especially for individuals who had vulnerabilities in healthcare use and financial sustainment. Emerging data show that the health and economic impacts of COVID-19 are being disproportionally borne by older adults who are socially vulnerable (Calderón-Larrañaga *et al*., 2020; Ayalon *et al*., 2021). As noted by prior studies (Scheppers *et al*., 2006; Wallace *et al*., 2013), coexisted unmet medical needs and financial hardships can exacerbate the self-efficacy of old persons and stimulate negative affections afterward. Positive aging attitudes, on the contrary, might promote the sense of capability among the vulnerable elderly and partially compensate for their positive cognitions (Moody, 2006). Particularly in the COVID-19 context, positive views regarding aging have direct and indirect associations with emotional well-being. More detailed, whereas ageism information is more likely to attack old persons with more significant vulnerabilities, positive aging attitudes help against such information and reduce negative ruminations (Tarazona-Santabalbina *et al*., 2021). Besides, older adults with confidence in their aging process are less likely to consider the accumulative risks of vulnerabilities as threats to their future lives (Swift *et al*., 2017), thus lowering the severity of negative emotions regarding current vulnerabilities. Thus, we propose that encouraging positive aging attitudes is essential for public health communication programs to promote emotional well-being among vulnerable older adults during adverse events.

### Strengths and limitations

This study reveals the latent classes of vulnerability for older adults in COVID-19 times and examines the relationships between vulnerability type, positive aging attitudes, and emotional responses. The findings of this study shed light on the detection of most vulnerable older adults and reinforce emotional well-being promotion strategies in a pandemic context. However, several limitations should be acknowledged. First, older adults that had been infected with the disease were underrepresented in this study. This study reported that 1.4% of respondents were diagnosed with the disease, whereas the prevalence may be much higher in the older adult population. Second, based on cross-sectional data, this study cannot infer causality, although it seems plausible in the temporal sequence vulnerability and aging attitudes first, and emotional responses being the outcomes. Third, there might be some confounding that was not controlled. For instance, we explained the moderation effect of positive aging attitudes with confrontation to ageism information. However, in the current study, whether and how the older adults were exposed to the ageism information was unknown. Despite these limitations, this study is one of the first studies to examine older adults’ emotional well-being with a vulnerability perspective. The findings of this study implicate emotional well-being promotion strategies in pandemic contexts. We propose that older adults’ emotional well-being should not be neglected. They deserve the support of prevention and intervention strategies, particularly when they have vulnerabilities in healthcare use and financial sustainment.

### Conclusion

We found older adults presented three distinct subclasses regarding their vulnerability in the COVID-19 times and about 28% of them experienced delayed healthcare needs solely or together with financial difficulties. Whereas older adults from different vulnerability types have no significant differences in their positive emotional responses, those with greater vulnerabilities are likely to have more negative emotions. Moreover, positive aging attitudes benefit older adults’ emotional well-being in the COVID-19 context, especially for individuals with vulnerabilities in both healthcare use and financial sustainment. Thus, we suggest public health communications and professional psychological interventions for older adults with females, BLOAC, and those aged below 65, uncoupled, less educated, and with ADL difficulties get prioritized.

## Data Availability

Data in this study was from Health and Retirement Survey (HRS). Processing code of this study is available by mailing to Dr. MQ Fu

https://hrsonline.isr.umich.edu/

## Description of authors’ roles

Mingqi Fu carried out the statistical analysis and drafted the manuscript. Jing Guo assisted with writing the article. Xi Chen supervised the data computation. Qilin Zhang designed the study and revised the manuscript.

## Acknowledgements

This study would like to thank the Health and Retirement Survey (HRS) group from University of Michigan for conducting this survey and sharing the data.

