## Supplementary material for "American older adults in COVID-19 Times:Vulnerability types, aging attitudes and emotional responses": Table S1

**TableS1. Comparing Models with Different Latent Classes: Fit indices (No. of Obs = 2003).**

| **No. of Groups** | **loglikelihood** | **AIC** | **BIC** | **ssaBIC** | **Entropy** | **LMR** | | **BLRT** | | **Proportion in class** | | | |
| --- | --- | --- | --- | --- | --- | --- | --- | --- | --- | --- | --- | --- | --- |
|  |  |  |  |  |  | 2LL | P | 2LL | P | 1 | 2 | 3 | 4 |
| **1** | -9828.10 | 19690.21 | 19785.45 | 19731.44 |  |  |  |  |  |  |  |  |  |
| **2** | -9315.25 | 18700.49 | 18896.57 | 18785.38 | 0.80 | 1018.27 | 0.00 | 1025.72 | 0.00 | 0.24 | 0.76 |  |  |
| **3** | -9223.31 | 18552.62 | 18849.55 | 18681.16 | 0.80 | 182.54 | 0.00 | 183.87 | 0.00 | 0.19 | 0.09 | 0.72 |  |
| **4** | -9167.00 | 18476.00 | 18873.77 | 18648.20 | 0.69 | 111.80 | 0.28 | 112.61 | 0.00 | 0.06 | 0.59 | 0.19 | 0.15 |

**Notes.**

**ssaBIC**: Sample-Size Adjusted BIC (n* = (n + 2) / 24); **LMR**: LO-MENDELL-RUBIN Test; **BLRT**: Bootstrapped Likelihood Ratio Test.

**Selection Criteria**: Model selection starts from one latent class and should stop if a) the AIC/BIC/ssaBIC begins to grow with another new group added; or b) the p value for LMR/BLRT turned nonsignificant with another new group added (p>0.05). Model with an Entropy of 0.8 or over is acceptable.
